# SARS-CoV-2 PCR cycle threshold at hospital admission associated with Patient Mortality

**DOI:** 10.1101/2020.09.16.20195941

**Authors:** Jui Choudhuri, Jamal Carter, Randin Nelson, Karin Skalina, Marika Osterbur-Badhey, Andrew Johnson, Doctor Goldstein, Monika Paroder, James Szymanski

**Affiliations:** Department of Pathology, Montefiore Medical Center, Albert Einstein College of Medicine, Bronx, N.Y; Department of Medicine, Saint Vincent’s Medical Center, Bridgeport CT USA

**Keywords:** COVID-19, Coronavirus, Coronavirus 2019, PCR cycle threshold

## Abstract

**Background:** Severe acute respiratory syndrome coronavirus 2 (SARS-CoV-2) cycle threshold (Ct) has been suggested as an approximate measure of initial viral burden. The relationship of initial Ct at hospitalization and patient mortality has not been thoroughly investigated.

**Methods and findings:** We conducted a retrospective study of SARS-CoV-2 positive, hospitalized patients from 3/26/2020 to 8/5/2020 who had SARS-CoV-2 Ct data within 48 hours of admission (n=1044). Only patients with complete survival data discharged (n=774) or died in hospital (n=270), were included in our analysis. Laboratory, demographic, and clinical data were extracted from electronic medical records. Multivariable logistic regression was applied to examine the relationship of patient mortality with Ct values while adjusting for established risk factors. Ct was analyzed as continuous variable and subdivided into quartiles to better illustrate its relationship with outcome. Cumulative incidence curves were created to assess whether there was a survival difference in the setting of the competing risks of death versus patient discharge.

Mean Ct at admission was higher for survivors (28.6, SD=5.8) compared to non-survivors (24.8, SD=6.0, P<0.001). Patients with lower Ct value on admission had higher odds ratio (0.91, CI 0.89-0.94, p<0.001) of in-hospital mortality after adjusting for age, gender, BMI, hypertension and diabetes. Patients with Ct values in 3rd Quartile (Ct 27.4-32.8) and 4th Quartile (Ct >32.9) had lower odds of in-hospital death (P<0.001). On comparing, Ct quartiles, mortality, BMI and GFR were significantly different (p<0.05) between the groups. The cumulative incidence of all-cause mortality and discharge was found to differ between Ct quartiles (Gray’s Test P<0.001).

**Conclusion:** SARS-CoV-2 Ct was found to be an independent predictor of patient mortality. However, further study is needed on how to best clinically utilize such information given the result variation due to specimen quality, phase of disease, and the limited discriminative ability of the test.

**AUTHOR SUMMARY:** Severe acute respiratory syndrome coronavirus 2 (SARS-CoV-2) pandemic has effected the entire world, with approximately 23 million affected till date. Clinicians, researcher and scientists are making all efforts to identify ways of diagnosis, predicting outcome and treatment modalities. The polymerase chain reaction (rT-PCR) technology, is the standard test being used for the diagnosis and it gives an additional value known as “cycle threshold” (Ct), which is the number of PCR cycles required to cross the designated threshold and termed patient as positive for the infection. This Ct value is inverse of the viral load in the patient and has been studied as indicator of outcome of infection. In this study we have analyzed the Ct value as a predictor for mortality and compared it between different age and gender. We found the Ct value significantly different between those who survived and those who died due to the disease. However proper utilization of the Ct value needs further studies to be utilized in the clinical setting and guide decision making.

## Introduction

Severe acute respiratory syndrome coronavirus 2 (SARS-CoV-2), the causative agent of COVID-19 is a novel betacoronavirus that first appeared in Wuhan, Hubei Province, China in late December 2019.(1) This virus has led to a global pandemic with over 23 million people infected worldwide with an overall mortality rate between 1.4% and 5%.(2, 3) It was declared a pandemic by the World Health Organization (WHO) on March 11, 2020.(4) While scientists, clinicians and researchers continue to grapple with the infection, the continuous in-flow of clinical data is helping to guide diagnostic, treatment and prognostic characteristics of the disease.

COVID-19 has wide-ranging clinical presentation, with 80% of cases being mild, 15% developing lower respiratory tract disease such as pneumonia, and less than 5% developing severe illness.(5) For those who progress to severe disease, the clinical course is insidious, with mild initial illness progressing to major complications in the second week.(5) The requirement for mechanical ventilation ranges from 18 to 33%, approximately 20% of hospitalized patients die of disease.(6-10) Multiple factors have been studied in association with disease severity, such as D-dimer, lymphopenia, obesity, hypertension and diabetes mellitus (DM) and there is still a continuous search for a reliable marker to predict disease aggressiveness.(11-16)

Real-time reverse-transcriptase (rRT-PCR) is a major modality of diagnosing infection. The cycle number (Ct) derived is the amplification required for the target viral gene to cross a threshold value and is inversely related to the viral load.(17) Kawase et al. have defined Ct as the cycle number when the sample fluorescence exceeds a set above the calculated background fluorescence.(18) There is limited literature exploring the association of Ct and disease mortality, and there has been variability in the findings.(10, 17, 19, 20) The objective of this study is to examine the relationship between SARS-CoV-2 cycle threshold at hospital admission and its relationship to patient outcome.

## Methodology

In this retrospective observational study we included all hospitalized patients at Montefiore Medical Center between 3/26/2020 and 8/5/2020 with a positive SARS-CoV-2 nasopharyngeal swab specimen on the Panther Fusion System (Hologic, Inc.) that was collected within 48 hours of admission. Subjects under the age of 18, current inpatients, those with initial SARS-CoV-2 testing on the Fusion system >48 hours after admission, or missing BMI were excluded from the analysis.(Figure 1)The patient information and data including test results and comorbidities were collected retrospectively from electronic medical records. Vital sign data included in the study, represented the first recorded vitals in the hospital records during admission and the biochemical and other parameters were indexed as the closest result within 48 hours to SARS-CoV-2 testing. The study was approved by the Albert Einstein College of Medicine Institutional Review Board.

**Figure 1.**
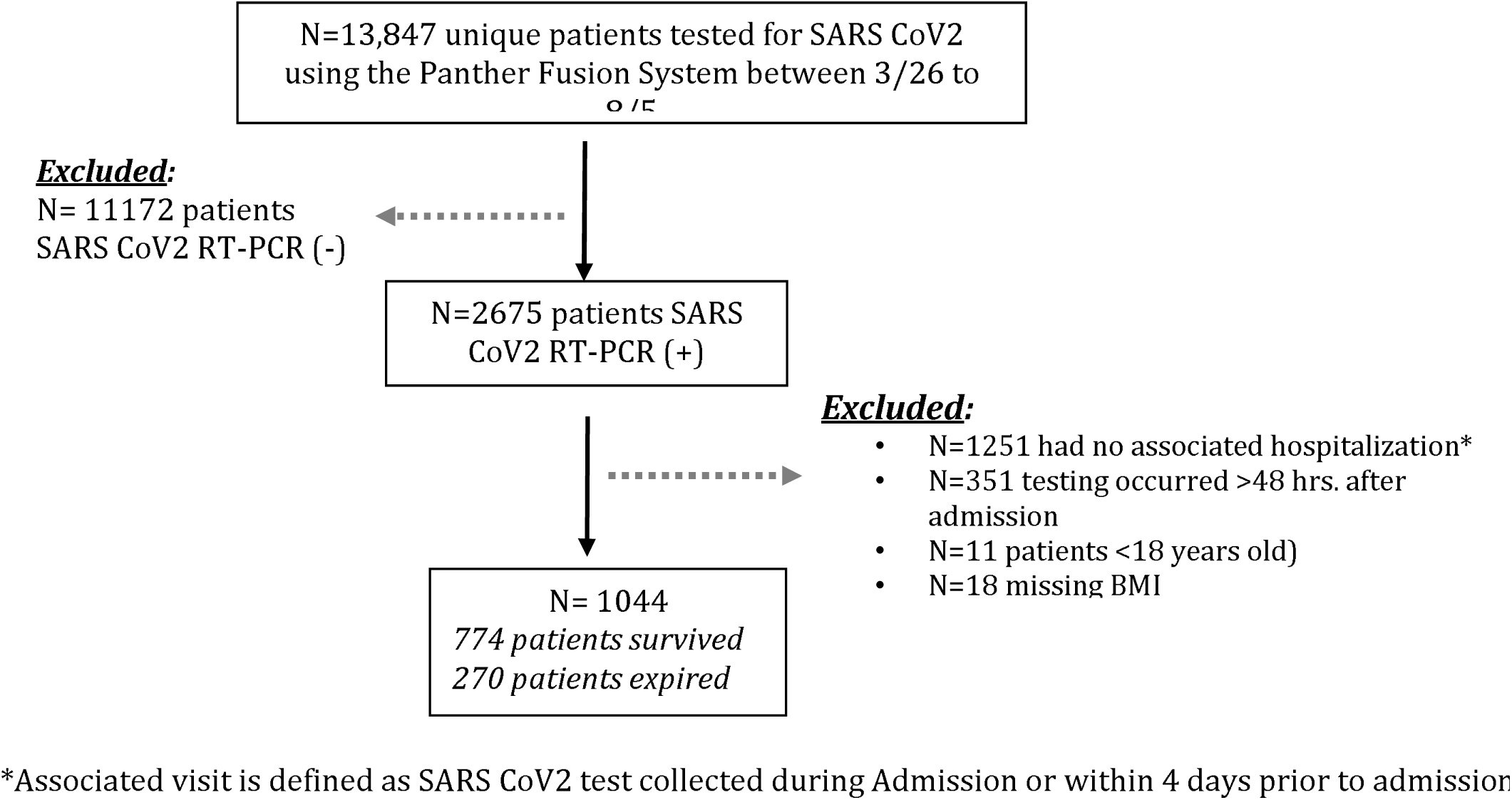
Study Cohort

Viral testing was performed on the Panther Fusion System (Hologic, Inc.) rRT-PCR platform, which has received Emergency Use Authorization with the Food and Drug Administration (FDA). All testing was performed according to the manufacturer’s instructions. The basic steps of the assay include sample lysis, nucleic acid capture, elution transfer, and multiplex RT-PCR where analytes are simultaneously amplified and detected. Results are reported as positive or negative depending on detectable amplification. The instrument also generates a Ct value. For the purpose of this study, Ct was divided into quartiles (Q1, Q2, Q3 and Q4) for grouping samples and studying the significance of viral load as an indirect marker. Low Ct corresponded to higher viral load.

### Statistical Analysis

Demographic and baseline group differences were assessed using chi-square tests, 2-sample Student t-tests, and for non-normally distributed data, the Mann-Whitney U test. The correlation between variables was assessed using the Kendall rank correlation method due to non-parametric data. The relationship between in-hospital mortality, cycle quartile, clinical risk factors, and biomarkers was modeled using logistic regression. Model covariate selection was based on literature review of identified SARS-CoV-2 survival risk factors. Model fit was assessed using the Hosmer and Lemeshow test which found p>0.05, thus not detecting evidence of poor fit. Multicollinearity was evaluated using variance inflation factors. Model discriminative ability was validated by calculating bootstrapped AUC. Cumulative incidence curves were created to assess whether there was a survival difference by cycle threshold quartile. This approach was used to account for the competing risks of in-hospital mortality versus patient discharge. A sensitivity analysis was carried out which assessed for patterns in missing data, study result sensitivity to covariate selection, and time period selected.(S1-S4) Analyses were performed using R version 3.6.2. A p-value <0.05 was considered statistically significant.

## RESULTS

A total of 1,044 patients met study inclusion criteria, of these 774 (74.1%) survived to discharge and 270 (25.9%) expired. Fig 1 In our cohort, 55.6% (580) patients were males and 44.4% (464) females with a mean age of 65.2 years (SD 15.37) and a mean BMI of 29.6 (SD 7.4). A history of hypertension (HTN) and Diabetes (DM) was present in 64.1% (629) and 40.8% (400), respectively. The majority of patients had group O blood-type, 46.5% (410), followed by group A blood-type, 31.6% (278).

Ct at admission was positively correlated with patient survival (r=0.22, p<0.001) (Figure 2). To better illustrate the magnitude of the effect, Ct was divided into quartiles (Q1, Q2, Q3 and Q4). The Q1 group consisted of cycle numbers <22.9, Q2 was cycle numbers between 23.0 and 27.3, Q3 was between 27.4 and 32.8 and Q4 was >32.9. It was noted that mortality was significantly different between the four groups, with highest mortality in those with lowest Ct (Q1=41.4%) and lowest in the group with highest Ct (Q4=13.2%) (p<0.001). Table 1. The association of cycle quartile and patient mortality held when stratified across age groups (Fisher’s exact test p<0.05 for all groups.) Fig 3. The cycle quartile was compared between age groups and gender. Fig 4. The cumulative incidence of hospital discharge varied across the cycle quartile (Gray’s Test P<0.001), as did the patient mortality (Gray’s test P<0.001). Fig 5.

**Table 1.**
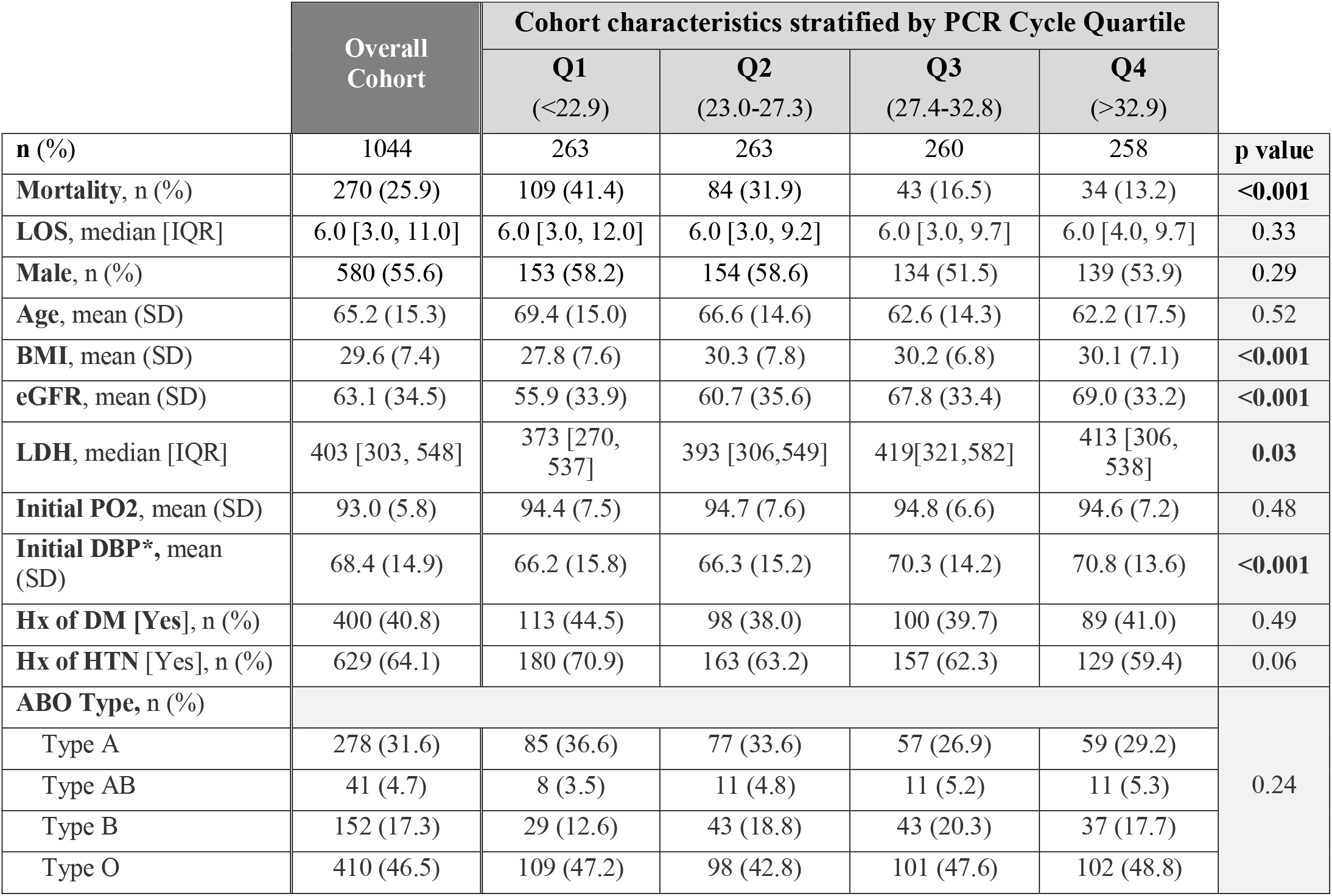
Study Characteristics by COVID-19 PCR Cycle Quartile

**Figure 2.**
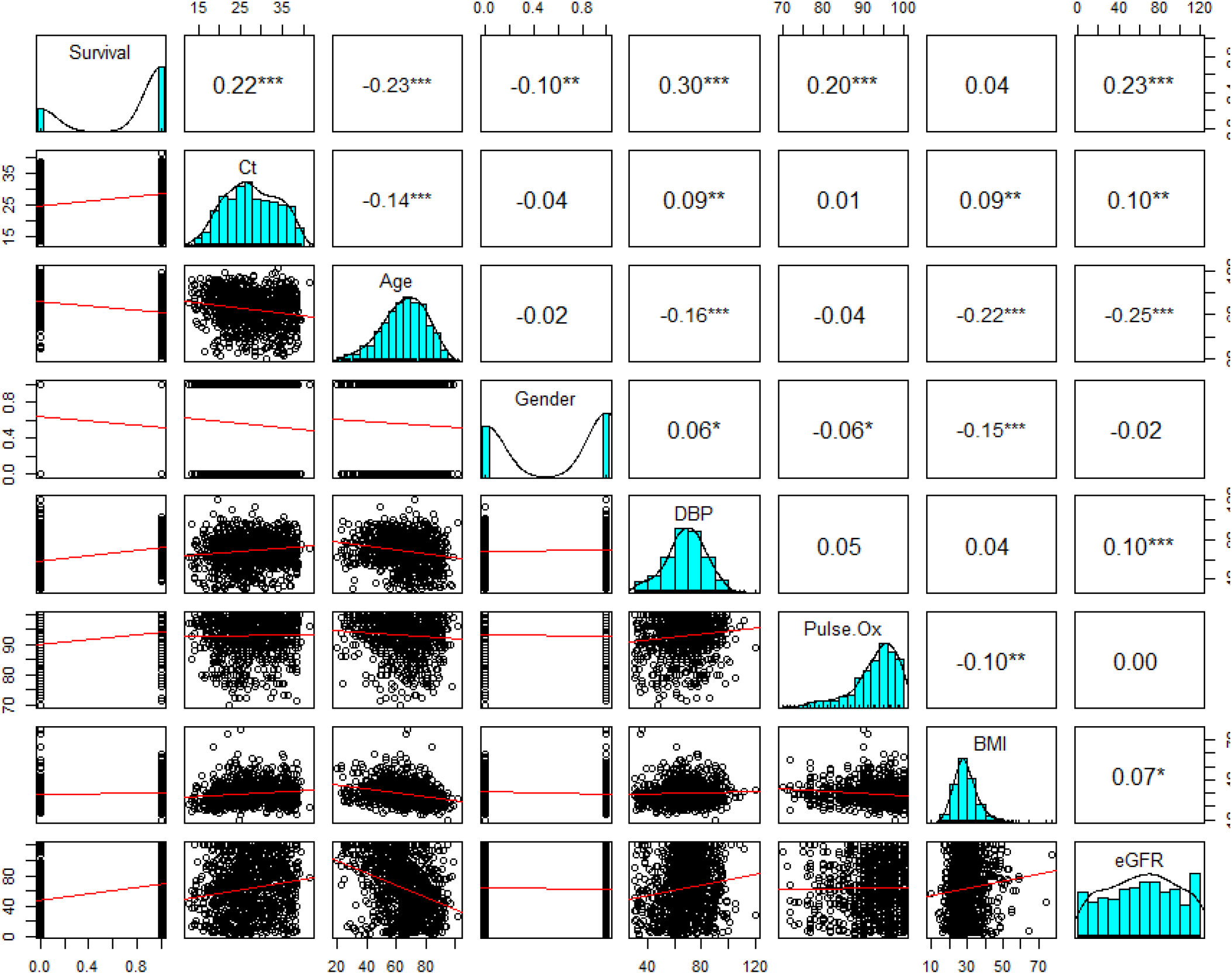
Correlation Matrix. On top is the (absolute) value of the Kendall correlation, the Asterix indicates degree of statistical significance: *p < 0.05, **p < 0.01, ***p < 0.001. On bottom, there are bivariate scatterplots, with a linear regression line.

**Figure 3.**
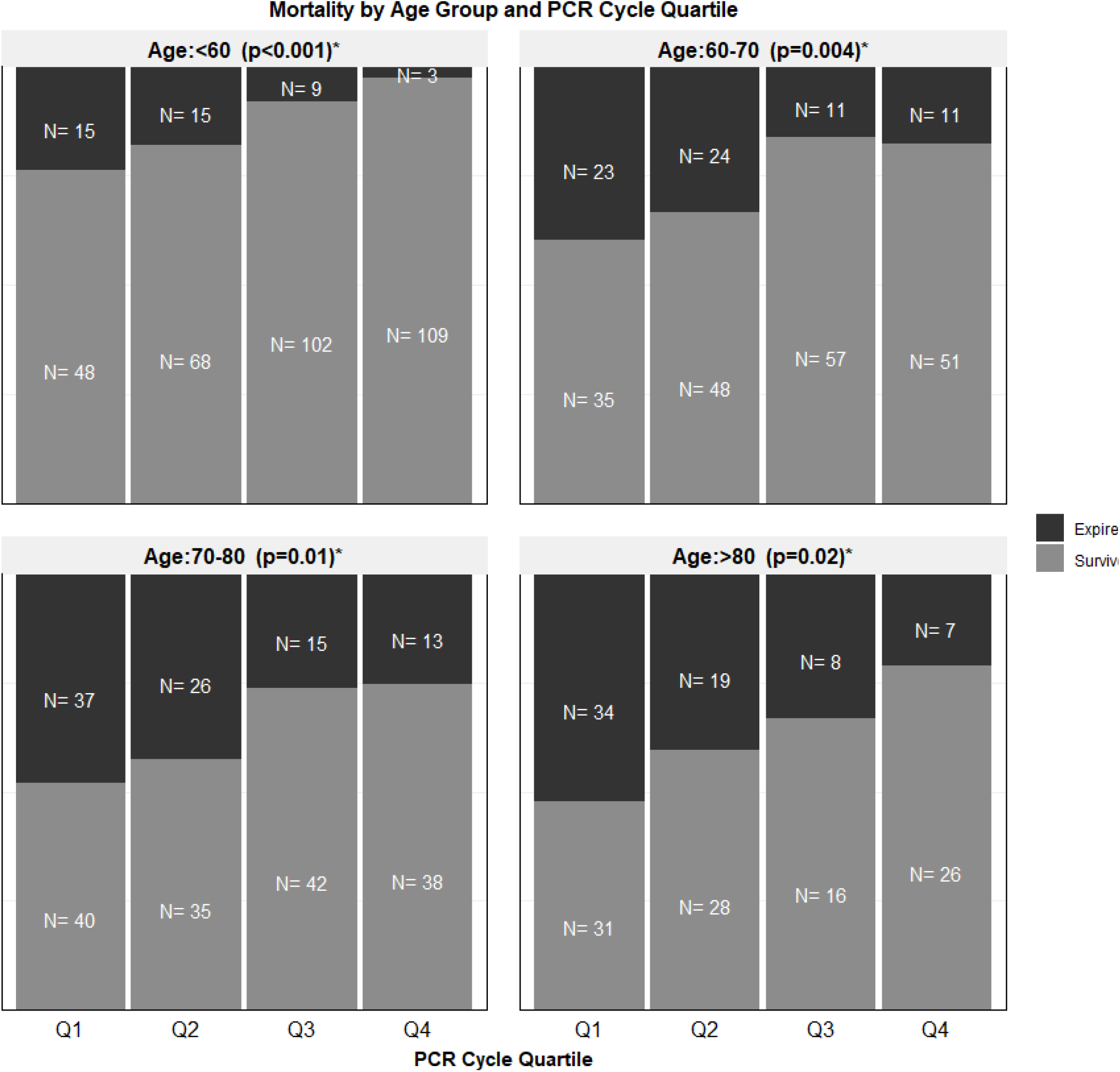
Stacked bar graph of Mortality by PCR Cycle Quartile and Age group. *Fisher’s exact test used.

**Figure 4.**
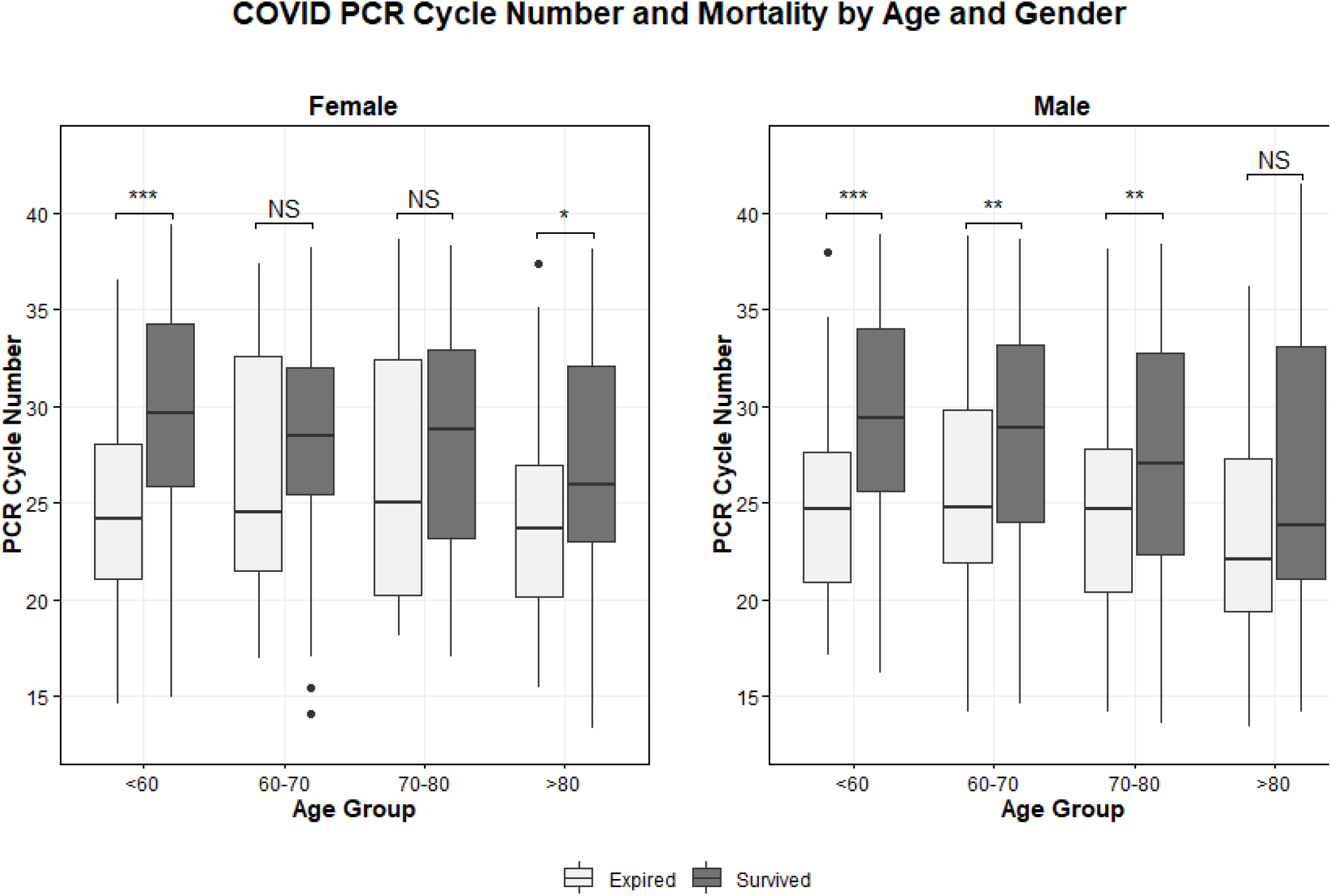
Cycle Number and Mortality by Age and Gender

**Figure 5.**
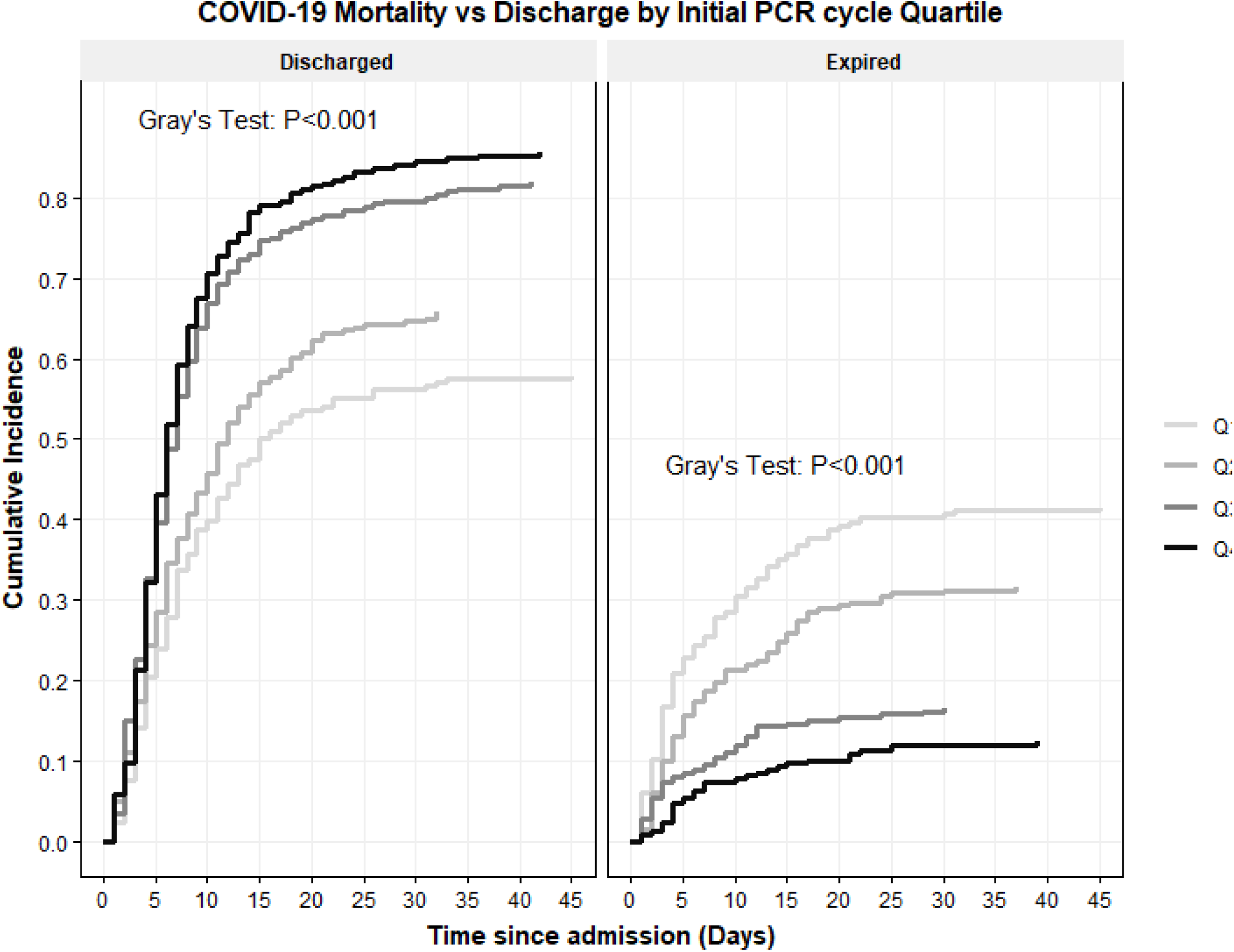
Cumulative Incidence function of COVID-19 Mortality vs Discharge by Initial COVID PCR Quartile

A multivariate logistic model was created describing the impact of cycle quartile, patient age, gender, BMI, and the comorbidities of hypertension and diabetes on patient survival. With the first cycle quartile (Q1) set as reference the adjusted odds of patient mortality decreased as cycle quartile increased, becoming significant in Q3 (aOR 0.34, CI 0.22-0.52, P<0.001) and Q4 (aOR 0.26, CI 0.16-0.42, p<0.001). Table 2. Male gender was found to confer a heightened risk of COVID-19 in-hospital mortality (aOR 1.88, CI 1.37-2.59, p<0.01). Likewise increased age, BMI, and a history of diabetes were all found to increase adjusted odds of in hospital mortality. Interestingly comorbid hypertension was found significant in univariable analysis but lost significance in the multivariable model.

**Table 2.** Logistic Regression table of In-Hospital Mortality

| In-Hospital Mortality |  | Expired | Survived | Odds ratio (univariable) | Odds ratio (multivariable) |
| --- | --- | --- | --- | --- | --- |
| **Age** | **Mean (SD)** | 72.4 (12.9) | 62.8 (15.4) | 1.05<br>(1.04-1.06, p<0.001) | 1.05<br>(1.04-1.06, p<0.001) |
| **Cycle Quartile** | **Q1** | 109 (40.4) | 154 (19.9) | - | - |
|  | **Q2** | 84 (31.1) | 179 (23.1) | 0.66<br>(0.46-0.95, p=0.024) | 0.68<br>(0.46-1.00, p=0.050) |
|  | **Q3** | 43 (15.9) | 217 (28.0) | 0.28<br>(0.18-0.42, p<0.001) | 0.34<br>(0.22-0.52, p<0.001) |
|  | **Q4** | 34 (12.6) | 224 (28.9) | 0.21<br>(0.14-0.33, p<0.001) | 0.26<br>(0.16-0.42, p<0.001) |
| **Gender** | **F** | 97 (35.9) | 367 (47.4) | - | - |
|  | **M** | 173 (64.1) | 407 (52.6) | 1.61<br>(1.21-2.15, p=0.001) | 1.88<br>(1.37-2.59, p<0.001) |
| **BMI** | **Mean (SD)** | 29.2 (8.2) | 29.8 (7.2) | 0.99<br>(0.97-1.01, p=0.301) | 1.03<br>(1.01-1.06, p=0.006) |
| **DM** | **No** | 139 (52.1) | 442 (61.9) | - | - |
|  | **Yes** | 128 (47.9) | 272 (38.1) | 1.50<br>(1.13-1.99, p=0.005) | 1.38<br>(1.00-1.90, p=0.048) |
| **HTN** | **No** | 74 (27.7) | 278 (38.9) | - | - |
|  | **Yes** | 193 (72.3) | 436 (61.1) | 1.66<br>(1.23-2.27, p=0.001) | 1.03<br>(0.72-1.47, p=0.868) |

The use of cycle threshold as a reported parameter predictive of inpatient mortality is limited by variation in sample collection, sample run variation, and the lack of a high-performing cutoff value Fig 6 and 7. The variation in optimal cutoff was assessed by bootstrapping with 1000 replications with ideal cutpoint determined using Youden’s index. The cutpoint which maximized Youden’s statistic was found to be 26 (CI 26-27). At this cutpoint, the testing set (out-of-bag) sensitivity was found to be 0.65 (CI 0.58-0.72) and specificity was 0.64 (CI 0.57 0.69). The area under the curve (AUC) was 0.68 (CI 0.63-0.71), thus the discriminative ability of the test to predict inpatient mortality was limited.

**Figure 6.**
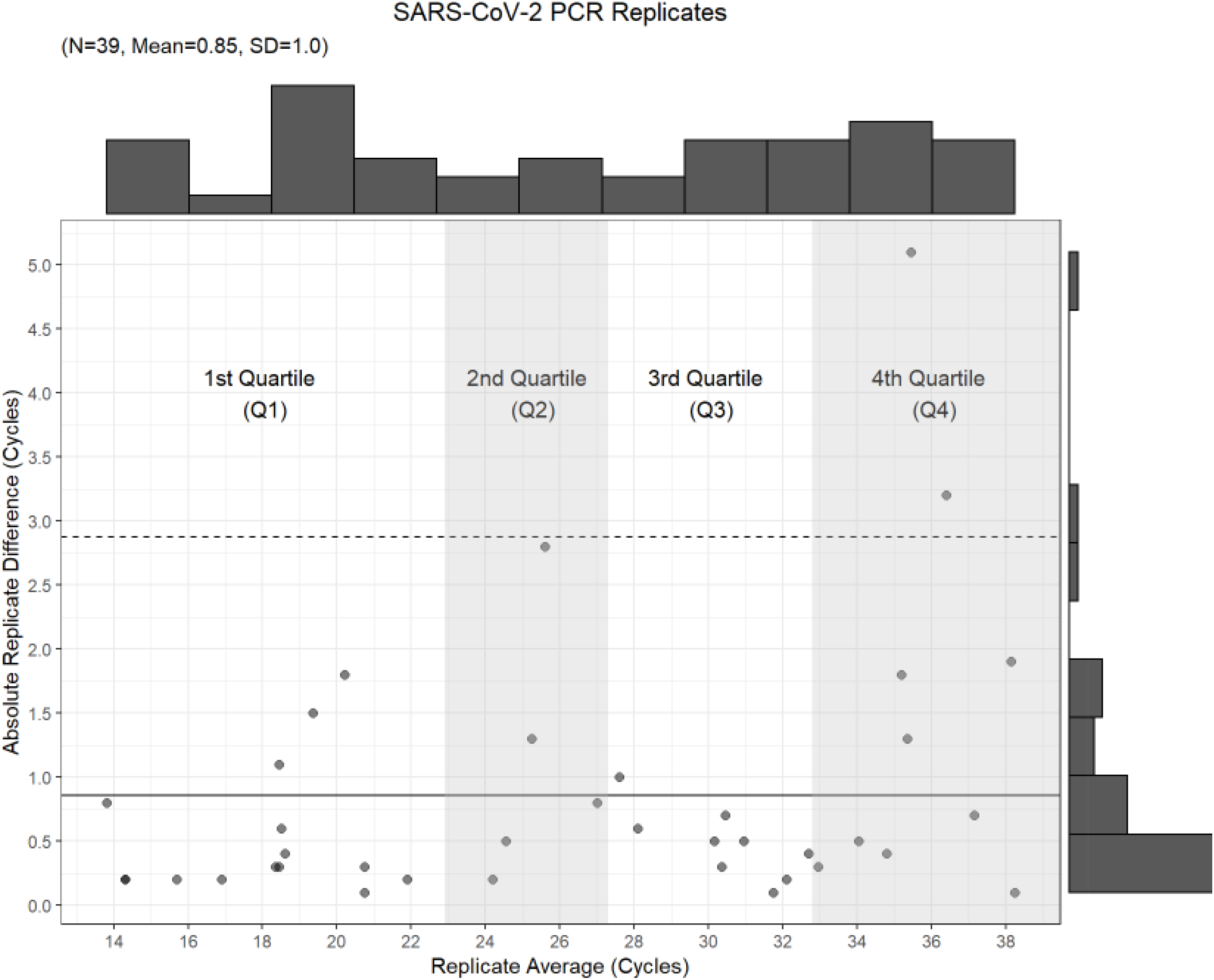
Scatterplot of averaged value of repeated samples versus absolute cycle number difference between replicates. The solid line indicates the mean absolute replicate difference, and the dashed line marks 2 standard deviations from the mean.

**Figure 7.**
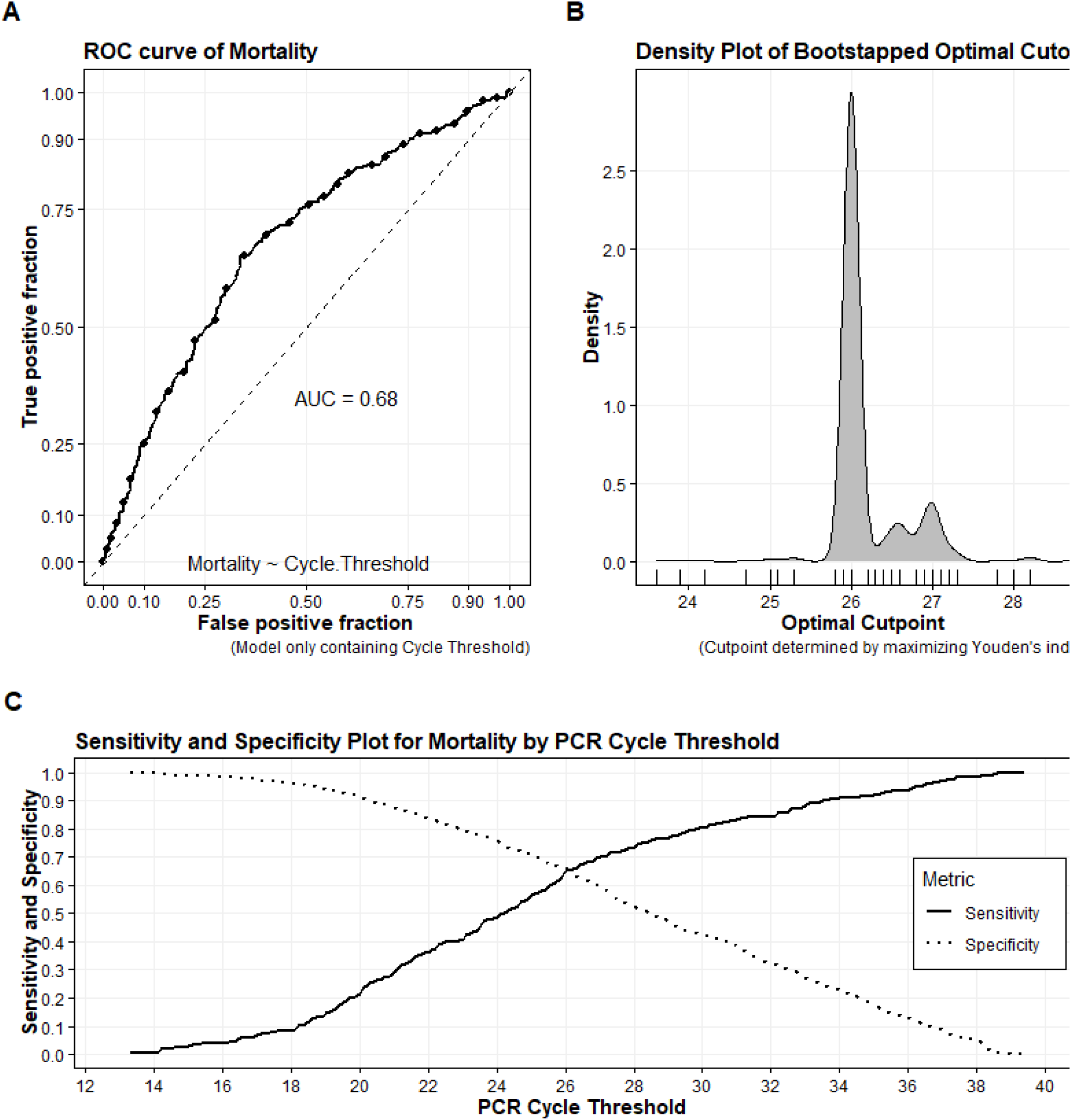
**(Panel A**): Receiver Operating characteristic (ROC) curve of Inpatient Mortality by Cycle Threshold value; (**Panel B**): Density plot of Bootstrapped Youden’s index cutoffs (1500 replications); (**Panel C**): Sensitivity and Specificity Plot of Mortality by PCR Threshold value

## DISCUSSION

COVID-19 has a highly-variable severity, thus identifying patients at risk of more aggressive disease is critical for triage and early management. While BMI, history of hypertension, and DM have been aggressively evaluated, there are recent studies which have looked at rRT-PCR Ct as an indicator of mortality or disease course.(10, 21) While it is difficult to utilize raw Ct values as a substitute for viral burden without the production of an appropriate standard curve or normalization to an internal housekeeping gene, attempts have been made to examine just this utility.(22) Previous studies which have looked at Ct or viral loads for disease aggressiveness have found differing results, which may be attributed to variability within the temporal course of infection.(10, 20, 23) Some studies have reported that peak load of the virus in upper respiratory tract specimens was expected during early stages of infection and in the pre-symptomatic stage, while others have found the peak to be approximately two weeks after the onset of symptoms even extending into the 3^rd^ or 4^th^ week of illness.(24-26)

Zheng et al. described increased viral load in respiratory samples in patients with a more aggressive disease course as compared to those with milder disease and therefore found that it could be used as a possible indicator of prognosis.(25) Similar results have been reported by Liu et al.(27) This correlation was not seen in relation to the viral load in stool samples.(25) Chen et al. reported that patients in the intensive care unit (ICU) continued to remain positive for COVID-19 infection longer than those who were not in the ICU.(28) Rao et al. in their systematic review of 18 studies reported an association between the viral load (or cycle number) and clinical outcome.(17) There was only one among 18 studies reviewed that looked at cycle number and mortality and found lower cycle number value associated with increased risk of mortality.(17, 29) Our results demonstrate a similar pattern as we found Ct values correlated inversely with mortality and low Ct increased the odds ratio for mortality compared to higher Ct.

According to Rao et al. 73% of the studies which looked at Ct in hospitalized patients found an association between Ct and disease aggressiveness, however, none of the studies which identified this association involved non-hospitalized patients.(17) Argyropoulos et al. in their research did report higher viral loads in non-hospitalized patients with an inverse relation between viral load and duration of symptoms and its severity.(20) The available literature suggests that recruitment of hospitalized patients might be a potential bias of analyzing the more severely ill among the overall infected population. Argyropoulos et al. did not find any association between viral load and length of hospital stay.(20) Our results also did not find any correlation with length of hospital stay for hospitalized patients.

Comparing our cohort of New York patients with findings by Magleby et al. it was noted that both showed a statistically significant difference between age groups and the Ct.(10) However, we also noted a difference related to male gender, which was not noted in their analysis.(10) Magleby et al. reported that patients with high viral loads were at increased risk of myocardial infarction and acute kidney injury.(10) We reached a similar inference regarding GFR and the Ct (viral load) showing that patient with high viral load had poorer GFR. Rao et al. found elevated LDH corresponded to low cycle numbers (higher viral load) and reported it as an important marker which showed consistent results in all the four studies in which it was included.(17) LDH was significantly different between patients of the different Ct quartiles in our study as well. Contrary to studies which have found ABO blood group as an indicator of disease severity, we were not able to identify such a correlation.(30)

Our study suggests that Ct can be used as an independent indicator of mortality--this corroborates previously reported data.(24) While Ct has been observed to be an important indicator of viral infection, it is influenced by the assay used and factors that can affect the amplification efficiency.(30) While studies like ours have helped identify independent markers which may in future help clinicians predict outcome, we are still cautious about its reliability for several important reasons. Firstly, most studies, including ours, have performed their analysis based on a single patient result. It is well understood that the testing is greatly limited by the quality of sample and its associated collection. This variability might be overcome through multiple patient sampling to improve the robustness of Ct as a tool. Additionally, some studies have reported contrary results with respect to viral load and disease outcome which might imply that the temporal relation of viral load and the Ct interpretation may depend on stage of infection. Literature on Ct is also limited by the fact that studies have primarily focused on hospitalized patients and the larger cohort of non-hospitalized patients and those with milder disease forms of infection have not been analyzed thoroughly enough.

While our study included a large and diverse cohort of patients due to its geographical location, some of the limitations included inability to analyze significance of race/ethnicity due to missing data from the medical records. This study could not explore viral load in non-hospitalized patients, and mild or asymptomatic patients were not initially swabbed due to hospital policy.

Ct-based risk stratification or interpretation is also limited by the absence of absolute or constant Ct cut-off values. The Ct value ranges can vary widely by platform and are impacted by amplicon length, target region, PCR cycling protocols, and other reaction conditions which alter the overall PCR efficiency even when the same target gene is amplified.(4) Adopting the Ct value for clinical judgement is limited by the scope of error due to multiple factors including interpretation by the examiner.(4) Additionally, since testing is primarily upon nasopharyngeal swabs there is much greater variability from one collection to another in addition to variability between patient disease states.(4) Therefore, even with our promising results, the clinical applications of our findings would require further investigation to determine the ultimate value of Ct interpretation.

## Conclusion

SARS-CoV-2 cycle threshold at admission was found to be an independent predictor of in-patient mortality. However, further study is needed on how to best clinically utilize such information given the result variation due to specimen quality, phase of disease, and the limited discriminative ability of the test.

## Data Availability

In medical electronic records

## SUPPLEMENTARY MATERIAL

**S1.**
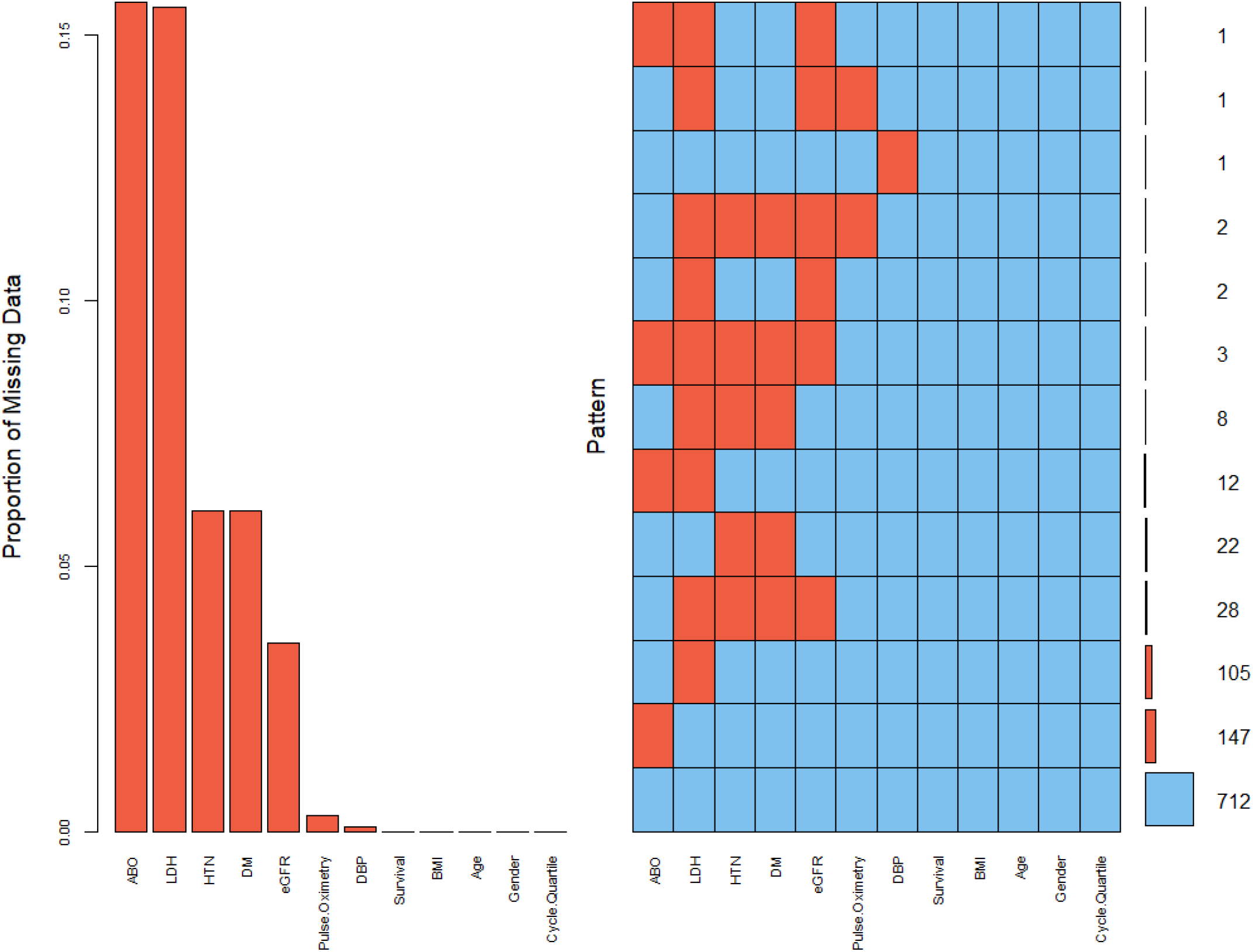
**(Left Panel)**: Barchart of the proportions of missing values. (**Right Panel)**: all existing combinations of missing values (Red) and non-missing values (Blue) with the pattern frequency represented by small horizontal bars with the number involved indicated.

**S2.** Sensitivity Analysis: examining the effect of selected time period on model results. Time period has been divided into quarters based on PCR testing totals

| *Predictors* | **Period 1** |  |  | **Period 2** |  |  | **Period 3** |  |  | **Period 4** |  |  |
| --- | --- | --- | --- | --- | --- | --- | --- | --- | --- | --- | --- | --- |
|  | *Odds Ratio*<br>*s* | *CI* | *p* | *Odds Ratio*<br>*s* | *CI* | *p* | *Odds Ratio*<br>*s* | *CI* | *p* | *Odds Ratio*<br>*s* | *CI* | *p* |
| **(Intercept)** | 0.13 | 0.01<br>–<br>1.60 | 0.11<br>1 | 0.04 | 0.00<br>–<br>0.81 | **0.036** | 0 | 0.00<br>–<br>0.05 | **<0.001** | 0.04 | 0.00<br>–<br>0.70 | **0.028** |
| **Age** | 1.03 | 1.01<br>–<br>1.05 | **0.003** | 1.05 | 1.02<br>–<br>1.08 | **<0.001** | 1.08 | 1.05<br>–<br>1.11 | **<0.001** | 1.06 | 1.03<br>–<br>1.09 | **<0.001** |
| **Gender [M]** | 1.18 | 0.67<br>–<br>2.10 | 0.56<br>8 | 2.67 | 1.36<br>–<br>5.26 | **0.005** | 2.55 | 1.30<br>–<br>5.00 | **0.006** | 1.71 | 0.89<br>–<br>3.30 | 0.108 |
| **BMI** | 1.01 | 0.97<br>–<br>1.05 | 0.71<br>3 | 1.04 | 0.99<br>–<br>1.09 | 0.12 | 1.08 | 1.03<br>–<br>1.14 | **0.003** | 1.03 | 0.99<br>–<br>1.07 | 0.102 |
| **Cycle Threshold** | 0.95 | 0.90<br>–<br>1.00 | 0.05<br>3 | 0.9 | 0.85<br>–<br>0.95 | **<0.001** | 0.91 | 0.86<br>–<br>0.96 | **0.001** | 0.9 | 0.85<br>–<br>0.94 | **<0.001** |
| **Observations** | 261 |  |  | 261 |  |  | 261 |  |  | 261 |  |  |

**S3.**
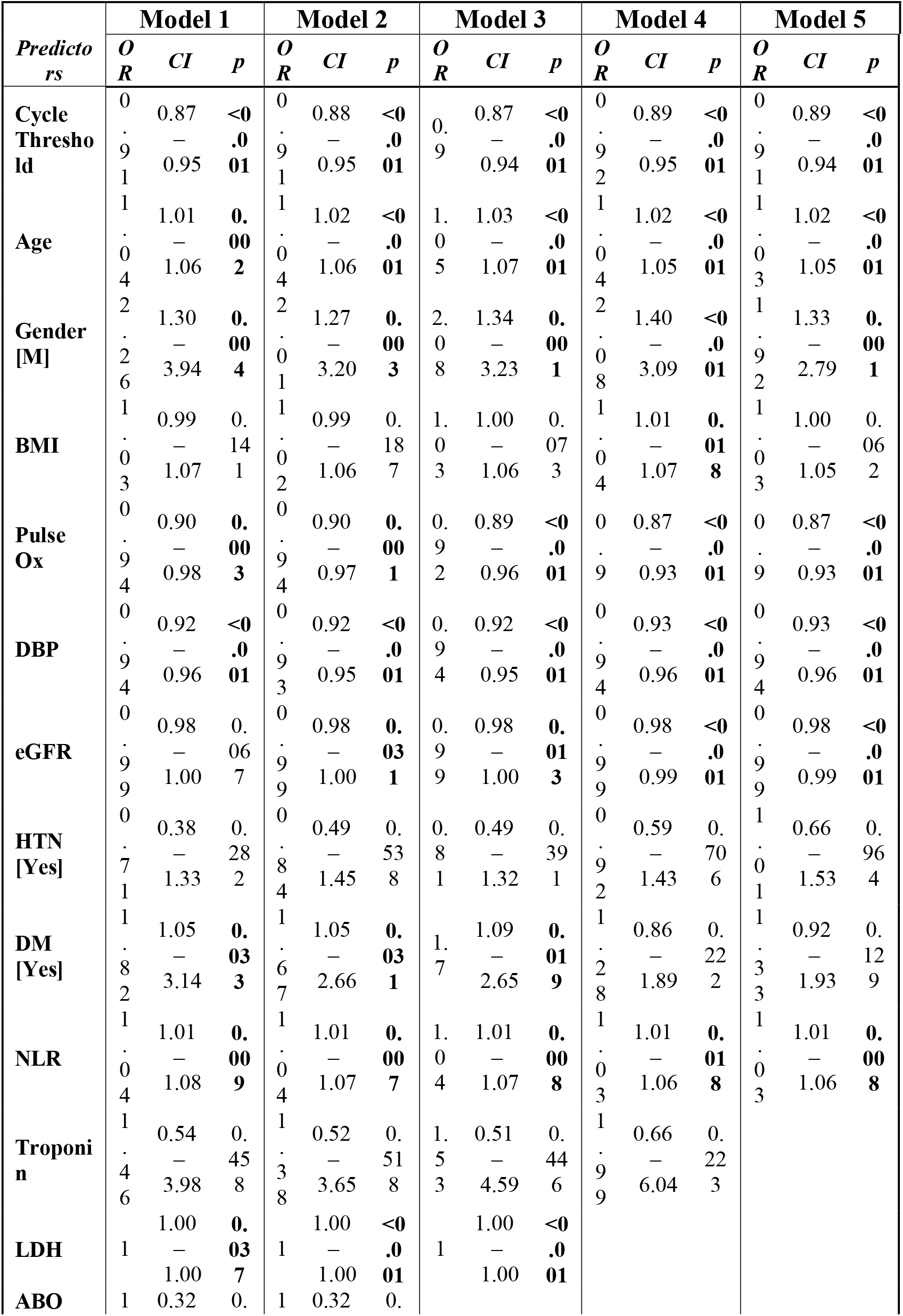
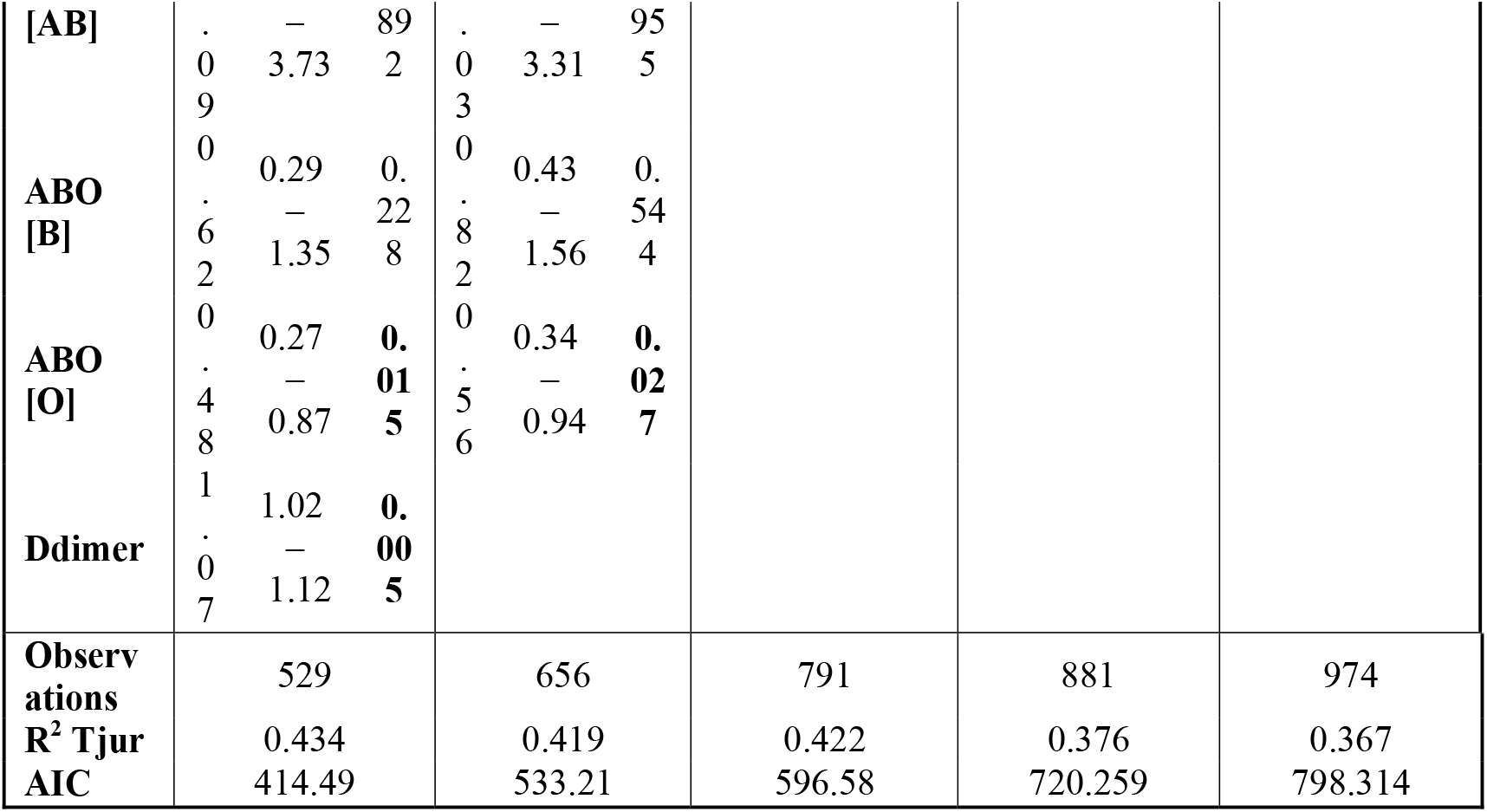
Sensitivity Analysis: examining the effect of covariate selection on study findings

**S4.** Sensitivity Analysis: examining the effect of covariate selection on study findings.

|  | Model 6 |  |  | Model 7 |  |  | Model 8 |  |  | Model 9 |  |  | Model 10 |  |  |
| --- | --- | --- | --- | --- | --- | --- | --- | --- | --- | --- | --- | --- | --- | --- | --- |
| *Predictors* | *OR* | *CI* | *p* | *OR* | *CI* | *p* | *OR* | *CI* | *p* | *OR* | *CI* | *p* | *OR* | *CI* | *p* |
| **Cycle Threshold** | 0.91 | 0.89 – 0.94 | <0.001 | 0.91 | 0.89 – 0.94 | <0.001 | 0.92 | 0.89 – 0.95 | <0.001 | 0.91 | 0.89 – 0.94 | <0.001 | 0.91 | 0.88 – 0.93 | <0.001 |
| **Age** | 1.03 | 1.02 – 1.05 | <0.001 | 1.03 | 1.01 – 1.04 | <0.001 | 1.03 | 1.02 – 1.05 | <0.001 | 1.04 | 1.03 – 1.05 | <0.001 | 1.05 | 1.04 – 1.06 | <0.001 |
| **Gender [M]** | 2.01 | 1.39 – 2.91 | <0.001 | 2.01 | 1.39 – 2.90 | <0.001 | 2.01 | 1.40 – 2.90 | <0.001 | 2.1 | 1.47 – 3.01 | <0.001 | 1.79 | 1.28 – 2.48 | 0.001 |
| **BMI** | 1.02 | 0.99 – 1.05 | 0.141 | 1.02 | 0.99 – 1.05 | 0.123 | 1.02 | 0.99 – 1.05 | 0.121 | 1.02 | 1.00 – 1.05 | 0.114 | 1.02 | 1.00 – 1.04 | 0.072 |
| **Pulse Ox** | 0.89 | 0.87 – 0.92 | <0.001 | 0.89 | 0.87 – 0.92 | <0.001 | 0.89 | 0.87 – 0.92 | <0.001 | 0.89 | 0.87 – 0.92 | <0.001 | 0.89 | 0.87 – 0.92 | <0.001 |
| **DBP** | 0.94 | 0.93 – 0.96 | <0.001 | 0.94 | 0.93 – 0.96 | <0.001 | 0.94 | 0.93 – 0.95 | <0.001 | 0.94 | 0.93 – 0.95 | <0.001 |  |  |  |
| **eGFR** | 0.99 | 0.98 – 0.99 | <0.001 | 0.99 | 0.98 – 0.99 | <0.001 | 0.99 | 0.98 – 0.99 | <0.001 |  |  |  |  |  |  |
| **HTN [Yes]** | 0.99 | 0.66 – 1.50 | 0.971 | 1.07 | 0.72 – 1.60 | 0.736 |  |  |  |  |  |  |  |  |  |
| **DM [Yes]** | 1.31 | 0.91 – 1.90 | 0.146 |  |  |  |  |  |  |  |  |  |  |  |  |
| **Observations** | 976 |  |  | 976 |  |  | 1006 |  |  | 1040 |  |  | 1041 |  |  |
| **R<sup>2</sup> Tjur** | 0.36 |  |  | 0.358 |  |  | 0.355 |  |  | 0.341 |  |  | 0.225 |  |  |
| **AIC** | 808.416 |  |  | 808.527 |  |  | 825.485 |  |  | 857.24 |  |  | 968.337 |  |  |

